# Rucaparib blocks SARS-CoV-2 virus binding to cells and interleukin-6 release in a model of COVID-19

**DOI:** 10.1101/2022.06.30.22277079

**Authors:** Henrietta Papp, Judit Bóvári-Biri, Krisztina Bánfai, Péter Juhász, Mohamed Mahdi, Lilian Cristina Russo, Dávid Bajusz, Adrienn Sipos, László Petri, Ágnes Kemény, Mónika Madai, Anett Kuczmog, Gyula Batta, Orsolya Mózner, Dorottya Vaskó, Edit Hirsch, Péter Bohus, Gábor Méhes, József Tőzsér, Nicola J. Curtin, Zsuzsanna Helyes, Attila Tóth, Nicolas C. Hoch, Ferenc Jakab, György M. Keserű, Judit E. Pongrácz, Péter Bai

## Abstract

Coronavirus disease 2019 (COVID-19), caused by SARS-CoV-2 virus, is a major global health challenge, as there is no efficient treatment for the moderate to severe disease. ADP-ribosylation events are involved in regulating the life cycle of coronaviruses and the inflammatory reactions of the host, hence we assessed the repurposing of registered PARP inhibitors for the treatment of COVID-19. We detected high levels of oxidative stress and strong PARylation in all cell types in the lungs of COVID-19 patients. Interestingly, rucaparib, unlike other PARP inhibitors, reduced SARS-CoV-2 infection rate through binding to the conserved 493-498 amino acid region located in the spike-ACE2 interface in the spike protein and prevented viruses from binding to ACE2. In addition, the spike protein-induced overexpression of IL-6, a key cytokine in COVID-19, was inhibited by rucaparib at pharmacologically relevant concentrations. These findings build a case for repurposing rucaparib for treating COVID-19 disease.

---

Coronavirus disease 2019 (COVID-19), caused by SARS-CoV-2 virus infection, is a global health challenge. SARS-CoV-2 is an enveloped virus with an ssRNA+ genome belonging to the *Coronaviridae* family^1^. SARS-CoV-2 predominantly infects the upper airways that may then transfer to lower airways causing atypical lung inflammation ^1^. The virus uses the angiotensin-converting enzyme 2 (ACE2) as cellular receptor for entry into epithelial cells^1^. Although, 80% of the patients develop mild or no symptoms, 15% develop severe disease requiring oxygen support, and 5% develop critical illness. The inflammatory response during the disease strongly contributes to organ damage and critical illness. Severe COVID-19 remains an unmet medical need calling for novel therapeutic modalities.

PARPs are ADP-ribosyl transferase enzymes composed of 17 members in humans (PARP1-PARP16)^2^. PARP1, PARP2 and PARP3 can be activated by damaged DNA^2^ that is often the result of reactive oxygen species (ROS) production under inflammatory conditions^2^. PARP activation contributes to necrotic and apoptotic cell death, furthermore, PARP activation has pro-inflammatory properties^2^. Four small molecule pharmacological PARP inhibitors, olaparib, rucaparib, niraparib and talazoparib are FDA/EMA-approved for cancer therapy and fluzoparib and pamiparib are approved by the Chinese NMPA ^2^.

Poly(ADP-ribosyl)ation (PARylation) of SARS-CoV-2 proteins can limit virus infectivity; a protective measure of the host that can be countered by the virus macrodomain^3,4^. A recent report suggested that stenoparib, a PARP inhibitor of both classical PARPs and tankyrases, can block the replication of SARS-CoV-2^5^. COVID-19 is characterized by ROS production and PARP activation^6^ that can contribute to cell death and tissue damage^2^. Taken together, PARP inhibition may have a dual pharmacological effect in COVID-19 disease, by blocking both SARS-CoV-2 replication and suppressing the consequent immune reaction. We set out to investigate the opportunity for repurposing registered pharmacological PARP inhibitors for COVID-19.

We have observed oxidative stress, marked by 4-hydroxynonenal (4HNE) staining, and PARP activation, marked by PAR immunostaining, in the pneumocytes, endothelial cells and macrophages of the lung tissue of COVID-19 patients (**Fig1A**), highlighting a role for PARylation in COVID-19.

**Figure 1.**
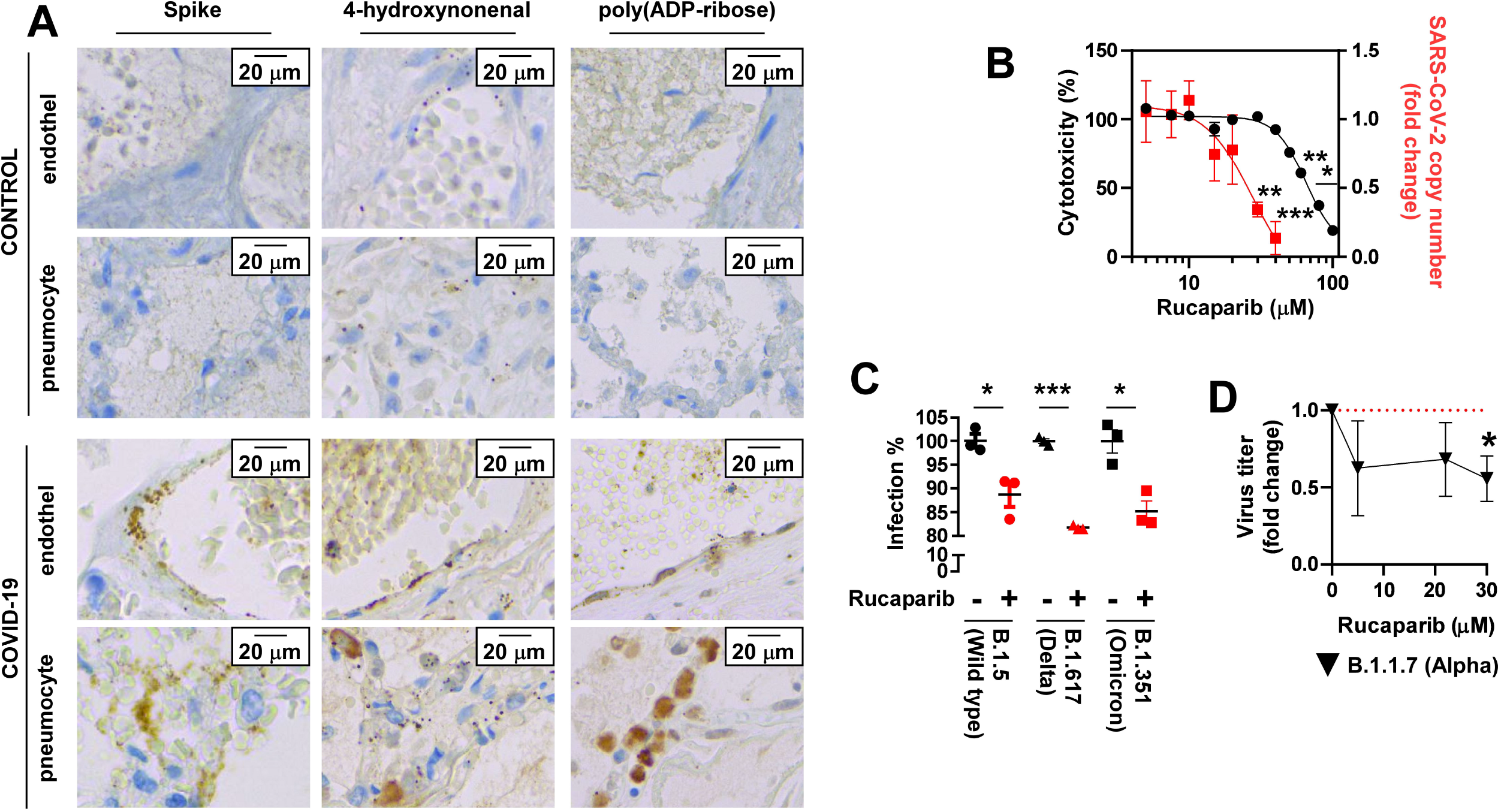
Rucaparib inhibits the binding of SARS-CoV-2 to the host cell. **(A)** In lungs of controls and patients died of COVID-19 SARS-CoV-2 spike protein, 4-hydroxynonenal and poly(ADP-ribose) immunohistochemistry was performed and evaluated. **(B)** The antiproliferative and toxic effects of rucaparib were tested on Vero E6 cells infected with of B.1.5 variant SARS-CoV-2. **(C)** Fluorescently-labelled pseudovirions were pre-treated with 35 µM rucaparib and pseudovirus uptake was assessed in HEK293T cells. **(D)** The alpha variant of SARS-CoV-2 was pretreated with rucaparib and was used to infect Vero E6 cells.

Next, we assessed three approved PARP inhibitors, rucaparib, talazoparib and olaparib in a cellular SARS-CoV-2 infection model in concentrations ranging up to 40 µM. Rucaparib inhibited the infection and proliferation of the B.1.5 variant of SARS-CoV-2 virus (original Wuhan variant with D614G mutation, IC_50_ = 27.5 µM, **Fig1B**), while talazoparib and olaparib had no effect (**FigS1A**). Importantly, rucaparib was not toxic at concentrations that profoundly inhibited viral proliferation (the IC_50_ value for toxicity was 64.8 µM **Fig1B**), hence, it is not direct toxicity that limits SARS-CoV-2 infection and proliferation.

The IC_50_ concentration for viral infection of rucaparib (27.5 µM corresponding to ∼8.7 mg/L) is much higher than that achieved at the recommended dose (600 mg BID, steady state level ∼2.4 mg/l^7^), furthermore, PARP inhibition of >90% is achieved using doses of 92 mg^8^. Therefore the impact on viral replication is unlikely to be due to PARP1 inhibition and may result from binding to other targets e.g. the spike protein. We tested this hypothesis in neutralization experiments. Rucaparib was preincubated with a pseudovirus bearing the spike protein of SARS-CoV-2 (wild-type, delta and the omicron variants) or the SARS-CoV-2 virus (alpha variant) and it inhibited virus uptake of all major variants of SARS-CoV-2 (**Fig. 1C-D**), suggesting that rucaparib abrogates the binding of the virus to host cell receptors. Rucaparib bound directly to the spike protein of SARS-CoV-2 (IC_50_ = 115±21.9 µM) in an *in vitro* assay, while no significant binding was detected for the other PARP inhibitors stenoparib and olaparib at concentrations up to 500 µM (**Fig. 2A**). We observed magnetisation transfer from the receptor-binding domain (RBD) of spike to rucaparib in saturation difference spectrum NMR (STD-NMR) experiments that verifies rucaparib binding to the RBD *in vitro* (**Fig2B**).

**Figure 2.**
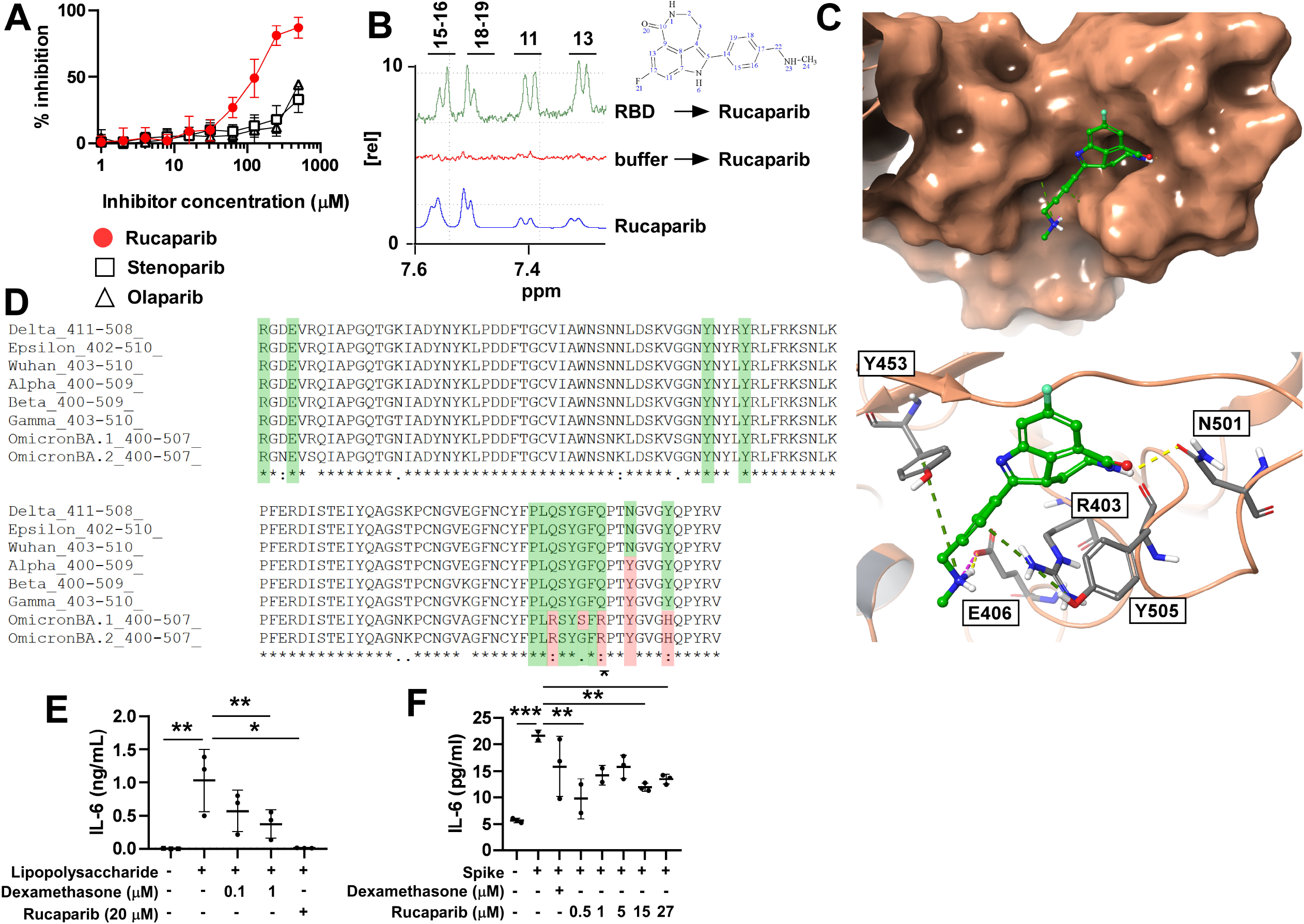
Rucaparib binds to the spike protein of SARS-CoV-2, and effectively inhibits IL-6 expression. **(A)** The indicated PARP inhibitors were tested to block the spike-ACE2 interaction in an *in vitro* binding assay. **(B)** STD-NMR spectrum of rucaparib alone and in complex with the receptor-binding domain of spike. **(C)** Predicted binding mode of rucaparib (green) at the protein-protein binding surface of the SARS-CoV2 RBD (fawn), with the main protein-ligand interactions highlighted as dashed lines (magenta: ionic interaction, yellow: H-bond, green: cation-pi interaction). **(D)** The sequence of the rucaparib binding site from the available variants were compared. **(E-F)** Human primary monocytes differentiated to macrophages were treated as indicated and IL-6 production was measured.

Next, we identified the putative binding site of rucaparib. A single binding hotspot was identified at the protein-protein interaction surface of all three variants (**FigS1B**), characterized by the residues 493-498, with further sidechains R403, E406, Y449, Y453, N501 and Y505 in the vicinity (**Fig2C**). The predicted binding mode of rucaparib is stabilized mainly by a strong ionic interaction between the negatively charged E406 sidechain and the terminal methylamine group of rucaparib. The latter is a positively charged and highly flexible moiety that is exclusive to this compound among the four PARP inhibitors tested in this study, which is in line with the exclusive on-target affinity of rucaparib. Additional interactions involve an H-bond between N501 and the NH group of the lactam unit, and back-to-back cation-pi interactions between Y453 and the methylamine group, as well as R403 and the phenyl ring. The STD-NMR shows magnetisation transfer (**Fig2B**) from RBD to H15, H16, H18 and H19 of the aromatic moiety of rucaparib that verifies binding and the validity of the *in silico* docking (**Fig2C, FigS1B**). Most amino acids responsible for rucaparib binding were conserved among SARS-CoV-2 variants. The omicron variant harbored the highest number of mutations at the binding site (**Fig2D**).

We assessed whether rucaparib targets the macrodomain of SARS-CoV-2 by applying a previously described model^3^. In agreement with previous results, Interferon-γ (IFNγ) treatment induced substantial cellular ADP-ribosylation that was reduced by ∼50% when the macrodomain of SARS-CoV-2 was overexpressed^3^. Rucaparib treatment at either 500 nM or 20 µM did not prevent this macrodomain-dependent reduction in IFN-induced ADP-ribosylation relative to control cells (**FigS1C**), suggesting that rucaparib does not inhibit the viral macrodomain at the concentrations tested. However, we observed a concentration-dependent effect of rucaparib on the overall induction of ADP-ribosylation by IFN-γ (**FigS1C**).

Next, we assessed the capacity of rucaparib to inhibit the expression of interleukin-6 (IL-6); a central cytokine in COVID-19^1^. In human macrophages, challenged by bacterial lipopolysaccharide (LPS), 20 µM rucaparib significantly reduced the expression of IL6, IL8 and IL10 at a similar or better efficacy as dexamethasone (**Fig2E, FigS2A**). Talazoparib (10 µM) and olaparib (20 µM) had no effect in this regard (data not shown). Subsequently, we induced IL-6 expression in macrophages with SARS-CoV-2 spike protein, and in this system rucaparib also blocked IL-6 overexpression (**Fig2F**) without affecting IL-1β and TNFα expression (data not shown). Finally, we assessed members of the Signal Transducer and Activator of Transcription (Stat) family of transcription factors (Stat1, 3, 5A, 5B), which are key mediators of interferon signaling and were implicated in COVID-19^9^. Stat1 and Stat3 were activated upon poly(I:C) or spike induction that was not inhibitedrucaparib at pharmacologically relevant concentrations (**FigS2B, C**). Interestingly, Stat5A and 5B activation was inhibited by spike, however, no statistically significant changes to Stat5A and 5B were elicited by rucaparib treatment (**FigS2C**).

We showed that rucaparib can bind to the spike protein of SARS-CoV-2 and, therefore inhibit the binding of SARS-CoV-2 to the ACE2 receptor and virus entry into host cells. These findings are similar to the reports on stenoparib that can inhibit SARS-CoV-2 proliferation in low concentrations^5^. Interestingly, olaparib and talazoparib had no effect on SARS-CoV-2 infection and proliferation. Most amino acids responsible for rucaparib binding are conserved among variants pointing out that rucaparib can be repurposed to block SARS-CoV-2 binding. Furthermore, the rucaparib binding site is a potential site for drug development to target the SARS-CoV-2 – ACE2 interaction.

Another important, and clinically relevant finding of the study is that rucaparib can efficiently block the expression of IL-6, the key interleukin involved in COVID-19-related inflammatory response^1^. In human monocyte-derived macrophages, rucaparib blunted both LPS or SARS-CoV-2 spike protein induced IL-6 overexpression. The lower rucaparib concentrations are pharmacologically relevant and are comparable to the achievable serum levels of rucaparib in humans^7,10^.

In the frame of this study, we assessed the applicability of pharmacological PARP inhibition in COVID-19 disease. We detected oxidative stress and PARylation in the lungs of COVID-19 patients similar to previous reports^6^. A large set of studies have provided evidence that PARP(1) activation is proinflammatory^2^. Several aspects of PARP-mediated inflammatory response have direct relevance to COVID-19-related inflammatory lung injury. Genetic or pharmacological PARP inhibition was protective in asthma, acute lung injury (burn, smoke inhalation, bacterial infection, etc.), ARDS, chronic obstructive pulmonary disease (COPD), lung fibrosis or ventilation-induced lung injury (reviewed in ^11^). Importantly, an overwhelming set of data point out that these findings can be translated to the human situation, for example, treatment with INO-1001; a PARP inhibitor, reduced serum IL-6 expression in humans^12^. The immunosuppressive effects of PARP inhibition in humans is further supported by a report^13^ showing that patients receiving PARP inhibitors as oncological treatment produced fewer neutralizing antibodies following SARS-CoV-2 vaccination as healthy volunteers.

Of note, there are other PARP-related events, relevant to COVID-19, which were not tested in our study. PARP overactivation can contribute to cell death and tissue damage^2,11^. Furthermore, PARP activation can strongly reduce cellular NAD+ levels^2^ that was implicated in the pathogenesis of COVID-19^6^. Several members of the PARP family, such as PARP9, PARP11 and PARP14 were also implicated in antiviral protection, including SARS-CoV-2^3,14-18^, often acting in different steps within the IFN signaling cascade. High doses of rucaparib may interfere with these PARP enzymes, as high dose rucaparib impaired IFN-induced ADP-ribosylation.

Taken together, rucaparib has a dual action in COVID-19, it can disrupt the binding of SARS-CoV-2 to ACE2 and it can target the inflammatory response. Although, the IC_50_ value of rucaparib in disrupting the SARS-CoV-2 binding is higher than the steady state levels on the dose approved for cancer therapy, rucaparib could potentially be applied as an aerosol in patients to achieve higher local concentrations so rucaparib can exert its dual effects in the lungs. Rucaparib, unlike the other clinically approved PARP inhibitors, was reported to continue to inhibit PARP1 for extended periods even after its removal^19,20^, further strengthening the case of its repurposing. Of note, the anti-inflammatory potential of rucaparib was comparable to dexamethasone, the standard of care in COVID-19, in the models used in this study. PARP inhibitors can potentially be synergistic with tocilizumab, and anti-IL-6 monoclonal antibody used against the cytokine storm, as well as with the anti-inflammatory drugs used in the therapy of COVID-19. These observations point towards the repurposing of rucaparib to combat COVID-19.

## Data Availability

All data produced are available online at https://figshare.com/s/6b25fd3d5de80ab3f51e

## Acknowledgement

The work was supported by grants from the NKFIH (K132623, K123975, K135150, K141142, TKP2021-EGA-10, TKP-2021-EGA-13, TKP2021-EGA-19, TKP2021-EGA-20, TKP2021-NVA-07). The Projects no. TKP2021-EGA-10, TKP-2021-EGA-13, TKP2021-EGA-19, TKP2021-EGA-20 were implemented with the support provided from the National Research, Development and Innovation Fund of Hungary, financed under the TKP2021-EGA funding scheme. Project no. TKP2021-NVA-07 has been implemented with the support provided from the National Research, Development and Innovation Fund of Hungary, financed under the TKP2021-NVA funding scheme. The POST-COVID2021-33 grant to PB was from the Hungarian Academy of Sciences. FAPESP grants 2018/18007-5 and 2020/05317-6 were to NH. The research was performed in collaboration with Cell and Tissue Culture Core Facility at the Szentagothai Research Centre of the University of Pecs. The work of D.B. was supported by the Janos Bolyai Research Scholarship of the Hungarian Academy of Sciences and the UNKP- 21-5 New National Excellence Program of the Ministry for Innovation and Technology. Prepared with the professional support of the Doctoral Student Scholarship Program of the Co-operative Doctoral Program of the Ministry of Innovation and Technology financed from the National Research, Development and Innovation Fund (to OM). The authors are grateful to Dr. Balazs Sarkadi (Institute of Enzymology, Research Centre for Natural Sciences, Budapest, Hungary) for his support and the critical revision of the manuscript.

## Conflict of interest

Rucaparib was a generous gift from Clovis Oncology (Boulder, CO, USA). Clovis had no impact on study design and the conclusions drawn.

Dr. Curtin is an inventor on patents WO 2005/012305 A2 and WO/2006/033006 with royalties paid to CRUK and Newcastle University, she gives her royalty share to charity and took no royalty in relation to this study.

Other authors declare no conflict of interest.

## Data availability

Primary data is available at https://figshare.com/s/6b25fd3d5de80ab3f51e (DOI: 10.6084/m9.figshare.19418957).

---

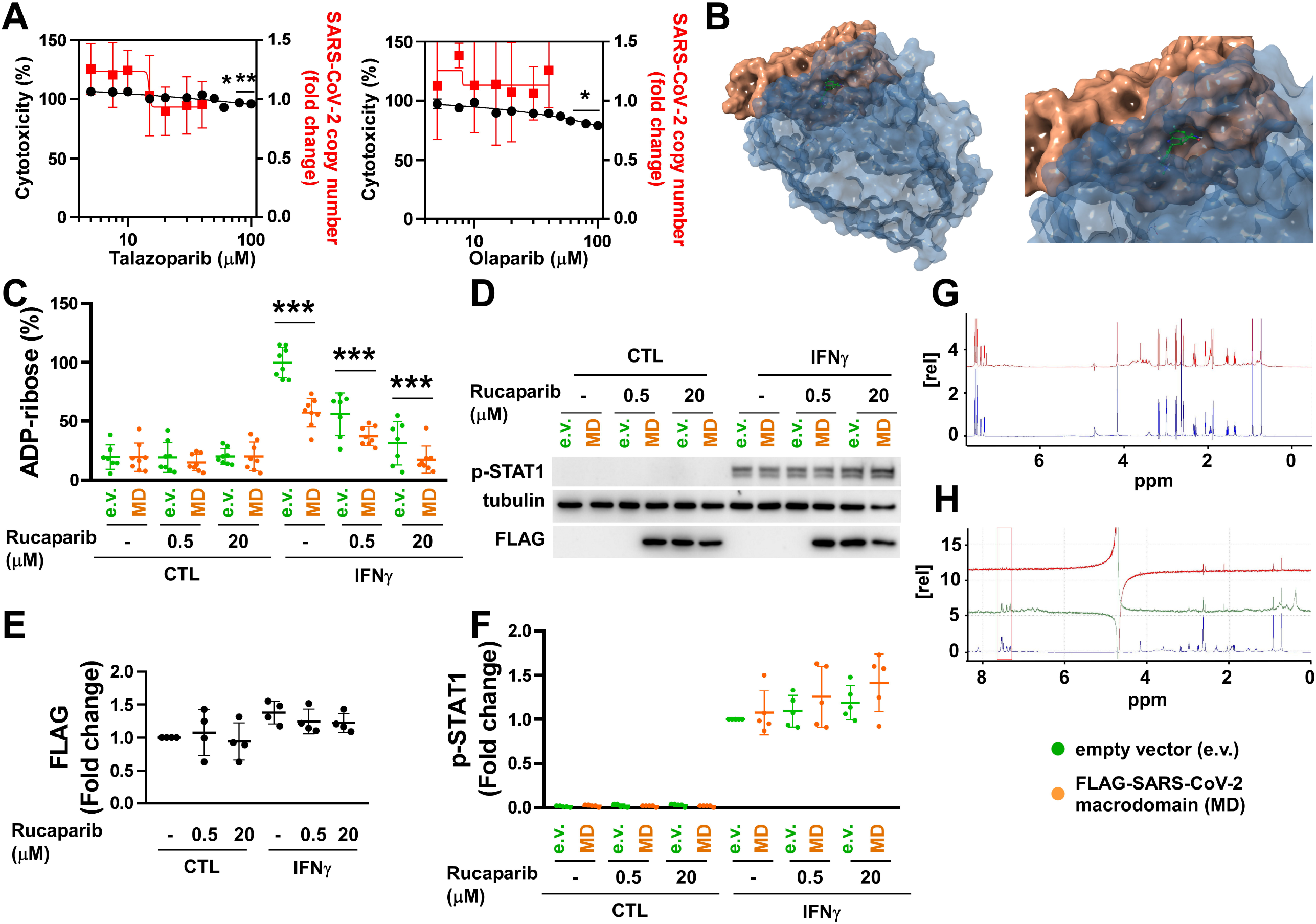

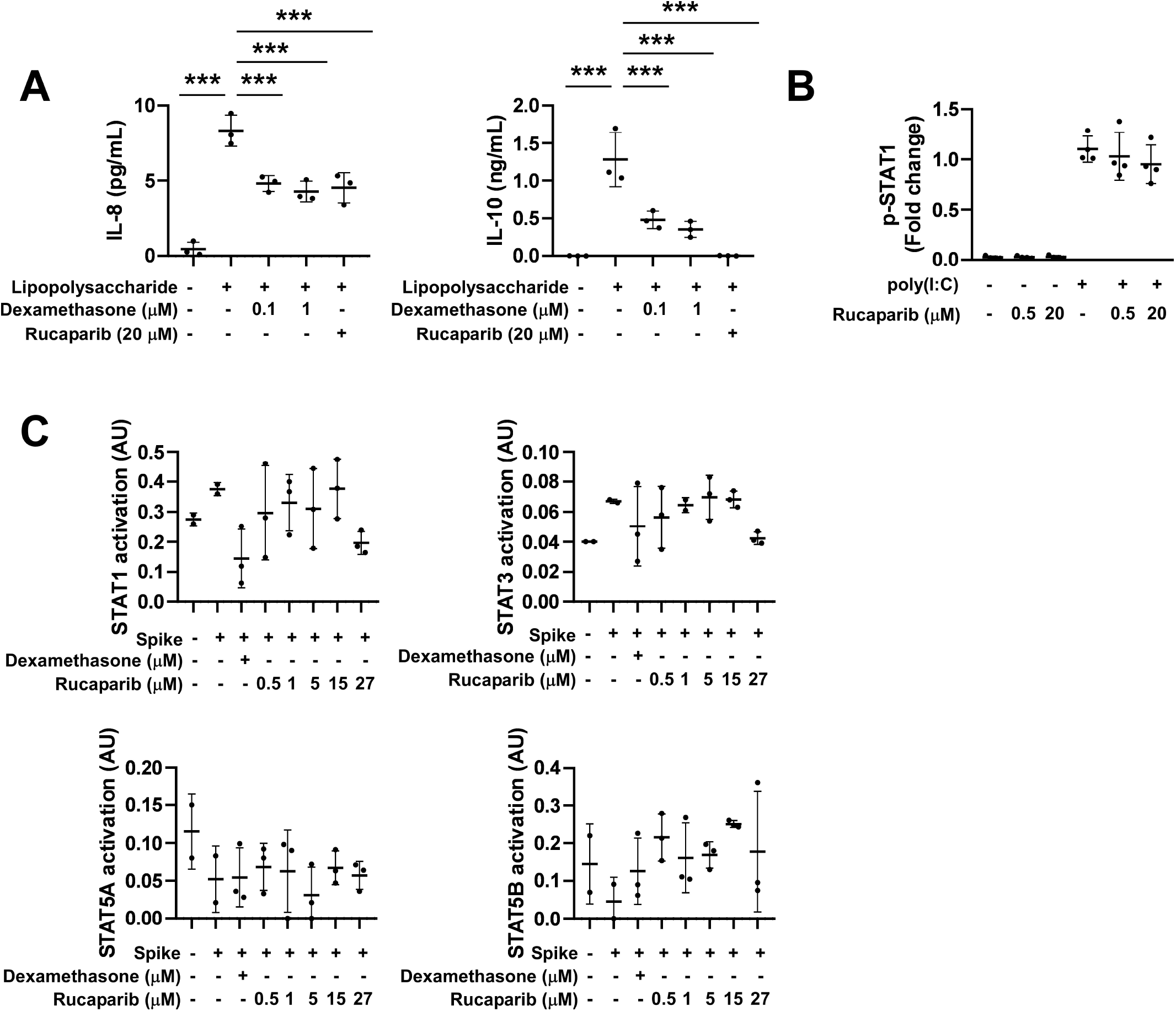

## Notes

### Author Declarations

Ethics Committee of the University of Debrecen, Faculty of Medicine gave ethical approval for this work (6043/2022).

