## Supplementary Methods and data for "Rucaparib blocks SARS-CoV-2 virus binding to cells and interleukin-6 release in a model of COVID-19"

*Chemicals*

Rucaparib was a generous gift from Dr. Thomas Harding (Clovis Oncology, Boulder, CO, USA), while Talazoparib and Olaparib was purchased from Sigma (St. Louis, MO, USA) and Stenoparib was from Selleckchem (Houston, TX, USA). LPS was from Sigma (St. Louis, MO, USA Cat. No: L4516), while the spike protein used to induce macrophages was from Merck (Kenilworth, NJ, USA, Cat. No.:AGX818).

*COVID-19 Patients*

Formalin-fixed, paraffin embedded (FFPE) blocks of patients was assessed who died of COVID-19 at Szent Erzsébet Hospital (Sátoraljaújhely, Hungary) between 30$^{th}$ November 2020 – 6$^{th}$ February 2021. Controls were patients died of non-COVID-19 causes. Controls were selected from patients died in the same period of non-COVID-19 and not lung-related causes at the clinical units of the University of Debrecen. Demographics of the patients are in **Supplementary Table 1**. We excluded patients with nosocomial infections in their records. The study was authorized by the local ethical board (6043/2022).

*Blood donors*

Monocytes were prepared from buffy coat of healthy donors. The study was authorized by the local ethical board (Reg. No. 938-2/2014/5200). Three-three donors were used in the LPS-induction and spike-induction experiments.

*Immunohistochemistry for poly(ADP-ribose), 4-hydroxynonenal (HNE) and spike protein*

Five micron thick sections were obtained from formalin-fixed, paraffin embedded blocks of COVID-19 patients. Immunohistochemistry was performed as in[1,2] with the conditions in **Supplementary Table 2**.

*Cell culture*

Vero E6 cells were cultured at 37 °C, 5% $CO_2$ in DMEM supplemented with 10% FBS and 1% penicillin-streptomycin.

Human embryonic kidney cells (HEK-293T) cells expressing ACE2 receptor and the TMPRSS2 protease (GeneCopoeia, Rockville, MD, USA) were grown in DMEM supplemented with 10 % FBS, and selection antibiotics (hygromycin 100 µg/mL, puromycin 1 µg/mL) and maintained in 37 C, 5% CO2 incubator.

A549 lung adenocarcinoma constitutively expressing FLAG-tagged SARS-CoV-2 Nsp3 macrodomain or its control empty vector[3] were grown in DMEM/high glucose media (Thermo) supplemented with 10% Fetal Bovine Serum (Thermo) and maintained at 37˚C in a humidified atmosphere containing 5% $CO_2$.

*Viruses*

SARS-CoV-2 B.1.5 (Accession ID: EPI_ISL_483637, with D614G mutation), B.1.1.7. (Accession ID: EPI_ISL_826270) and B.1.617.2 (Accession ID: EPI_ISL_9625925) variants were isolated at the National Laboratory of Virology (University of Pécs, Pécs, Hungary). Variants were verified by sequencing.

SARS-CoV-2 spike protein-pseudotyped lentivirions were produced by transfection of HEK-293T cells with the following plasmids: pLenti CMV GFP Puro expression vector, psPAX2 packaging plasmid (Addgene, Watertown, MA, USA), and pcDNA3.1 plasmids coding for either the Wuhan-Hu-1 prototypical, the 1.617 or the 1.351 SARS-CoV-2 spike variant (GenScript Biotech, Piscataway NJ, USA). Transfection was done in a 1:1:1 ratio using the polyethylenimine (PEI) method, and virion production was carried out as described previously [4]. Following collection and concentration of the pseudovirions, an enzyme-linked immunosorbent assay (ELISA)-based colorimetric reverse transcriptase (RT) assay (Roche Applied Science, Mannheim, Germany) was then used to detect the amount of RT in the virus samples, and transduction (infection) of cells was carried out using 4 ng RT-equivalent of the pseudovirions (MOI 0.2).

*Virus proliferation assay*

In a 96-well cell culture plate, 3x $10^5$ Vero E6 cells were seeded on the previous day of the infection. Treatment with the different PARP inhibitors (eucaparib, olaparib, talazoparib) from 40 – 2.5 µM were done at the same time with the infection of SARS-CoV-2 (Accession ID: EPI_ISL_483637, B.1.5 variant) at MOI 0.01 in a BSL-4 laboratory. After a 30 minutes of incubation in a cell culture $CO_2$ incubator, the mixture of the virus and the compound was replaced with only compound containing cell culture media (DMEM, 2% HI FBS, 1% PS). 48 hours post infection the supernatant was collected. RNA extraction was performed using the Monarch Total RNA miniprep kit (New England Biolabs, Ipswitch, MD, USA). Viral titer from the supernatant was determined using a QX200 AdutoDG Droplet- Digital PCR using the ddPCR Supermix (both from Bio-Rad, Hercules, CA, USA). The PCR was specific for the SARS-CoV-2 RdRp gene (forward primer: GTGARATGGTCATGTGTGGCGG; reverse: CARATGTTAAASACACTATTAGCATA, probe FAM-CAGGTGGAACCTCATCAGGAGATGC-BBQ). $IC_{50}$ was determined by performing a non-liner regression using the [Inhibitor] vs.

response variable slope (four parameters) macro of Graphpad Prism. For rucaparib the R2 value was $R^2_{infection}$=0.8176.

*Viability assays*

Cell viability was assessed in Vero E6 cells using the CellTiter-Glo assay (Promega, Madison, WI, USA). $IC_{50}$ was determined by performing a non-liner regression using the [Inhibitor] vs. response variable slope (four parameters) macro of Graphpad Prism. The $R^2$ value for the goodness of fit was $R^2_{viability}$=0.9776 for Rucaparib.

*Neutralization assay on Vero E6 cells*

8 x $10^5$ Vero E6 cells were seeded in a 48-well plate. The next day the plate was transferred to the BSL-4 laboratory. SARS-CoV-2 B.1.1.7 viral stocks were diluted to 0.01 MOI in DMEM (Lonza, Basel, Switzerland). Virus suspension (25 µl) was mixed with 25 µl rucaparib (60, 44 and 10 µM diluted in DMEM) and incubated for an hour at room temperature. After the incubation period 450 µl DMEM was added to the mixture then it was added to the cells. The cells were infected for 30 min at 37 °C. The SARS-CoV-2 and rucaparib mixture was then discarded and the cells were incubated for 2 days with DMEM supplemented with 2% HI FBS and 1% Pen-Strep. 48 hours post infection the supernatant was collected. RNA extraction was performed using the EXM 3000 nucleic acid isolation system (Zybio, Chongqing, China). Viral titer from the supernatant was determined using a QX200 AdutoDG Droplet- Digital PCR using the ddPCR Supermix (both from Bio-Rad, Hercules, CA, USA). The PCR was specific for the SARS-CoV-2 RdRp gene (forward primer: GTGARATGGTCATGTGTGGCGG; reverse: CARATGTTAAASACACTATTAGCATA, probe FAM-CAGGTGGAACCTCATCAGGAGATGC-BBQ).

*Neutralization assay on hybrid HEK293T cells*

The day before the experiment, HEK-293T cells expressing ACE2 receptor and the TMPRSS2 protease (GeneCopoeia, Rockville, MD, USA) were passaged into 48-well plate (30 000 cells/well) in DMEM containing 10 % FBS and 1 % penicillin-streptomycin. On the next day, 35 µM of rucaparib in DMSO was incubated with 4 ng/ul RT-equivalent of the pseudovirions for 30 minutes in DMEM medium without serum or antibiotics (50 µl total), in 37 C. For controls, equivalent amount of DMSO was added to the pseudovirions in the absence of rucaparib. The media on the cells were then removed, and the virus-inhibitor medium was complemented with 450 µl DMEM containing 10 % FBS, 1 % penicillin-streptomycin, and thereafter added to the wells (48-well plate). The cells were then incubated for 48 hours in 37 C, after which the media was removed from the wells, and the cells were collected in PBS (400 µl) for analysis of GFP fluorescence indicating transduction using flow cytometry (FACSCalibur, BD Bioscience,

Franklin Lakes NJ, USA), counting 3000 cells/sample. Measurement was carried out in triplicates. Cells were visually inspected for signs of cell death and signs of cell death was not observed in any of the experiments reported.

*Monocyte differentiation*

Peripheral blood mononuclear cells (PBMC) were isolated from three different buffy coat samples received from the Hungarian National Blood Transfusion Service. To separate mononuclear cells, we layered the diluted blood sample in 1:1 ratio with basal RPMI 1640 Medium (Lonza, Cat. No: 12-167F) onto the Lymphocyte Separation Medium 1077 (PromoCell, Cat. No: CC-44010.) and applied a centrifugation step (400 g, for 30 min, at room temperature with4/1 acceleration/deceleration). The layer of the mononuclear cells was transferred, washed with PBS (Lonza, Belgium, Cat. No: 17-516F) and centrifuged at 300 g for 10 min at room temperature. Following cell counting $5x10^5$ cells per well were seeded to 24-well plates (Greiner, Germany, Cat. No: 662160) in complete RPMI 1640 Medium supplemented with Fetal Bovine Serum (Biosera, USA, MO, Cat. No: FB-1350-500) Penicillin-Streptomycin (Lonza, Belgium, Cat. No: DE17-602E) and L-glutamine (Lonza, Belgium, BE-17-605E). To induce macrophage differentiation, we incubated the cells for 5 days at 37 ˚C, 5% $CO_2$ in a humidified atmosphere.

*Treatment of differentiated macrophages*

In the first part of the experiment, differentiated macrophages were treated in three parallels with the combination of 100 ng/ml LPS (Sigma, USA, MA, Cat. No: L4516) and three different PARP inhibitor molecules diluted in DMSO (Santa Cruz, USA, CA, Cat. No: sc-358801). A 24-hour treatment was applied using 20 µM concentration of Rucaparib and Olaparib and 10 µM concentration of Talazoparib. In all cases 0.1 % DMSO concentration was used as solvent control. The positive control was treated with 0.1 µM and 1 µM Dexamethasone (Teva, Hungary, Cat. No: OGYI-T-6071/03). Following incubation supernatants were collected for further investigation. Monocytes from three donors were accessed and were plotted. The vehicle was 0.01% DMSO.

In the second part of the experiment, three parallels of differentiated macrophage cells (three different donors, 30 000 cells/well) were pretreated for 48 hours with 20 nM spike protein (Merck, Germany, Cat. No: AGX818). Rucaparib was added after 48 hours in 5 different concentrations (500 nM, 1 µM, 5 µM, 15 µM, 27 µM) and incubated for 24 hours. The positive control was treated with 1 µM Dexamethasone (Teva, Hungary, Cat. No: OGYI-T-6071/03). Supernatants and cell pellets were collected for further assays. One representative donor is displayed in three biological replicates due to a large heterogeneity of the donor responses.

*Inflammatory cytokine concentration measurement using Luminex xMAP technology*

Luminex Multiplex Immunoassay was performed to determine the following cytokines/chemokine concentrations using customized Milliplex Human Cytokine/Chemokine/Growth Factor Panel A Magnetic Bead Panel (Cat. Nr. HCYTA-60K, Merck Millipore, Darmstadt, Germany): interleukin-10 (IL-10); IL-1β; IL-2; IL-6; IL-8; tumor necrosis factor-alpha (TNF-α). Following previous optimizations, the undiluted samples were tested in a blind-fashion and in duplicate. The experiment was performed according to the instructions of the manufacturer. Briefly, 25 µl volume of each sample, control, and the standard was added to a 96-well plate (provided by the kit) containing 25 µl of fluorescent-coded, capture antibody-coated beads. After the appropriate washing and incubation periods, biotinylated detection antibodies and streptavidin-PE were added to the plate. 150 µl volume of drive fluid was added to the wells after the last washing step, and the plate was incubated and read on the Luminex MagPix instrument. Five-PL regression curves were used to plot the standard curves for the analytes by the Belysa 1.1 (Merck KGaA, Darmstadt, Germany) software analyzing the bead median fluorescence intensity. Results are shown in pg/mg.

*Inflammatory cytokine concentration measurement using the Proquantum kit series*

In spike induction experiments IL-6 (A35575), TNFα (A35601) and IL-1β (A35574) were determined using the respective Proquantum assay kits (ThermoFisher, Waltham, MA, USA) according to the instructions of the manufacturer.

*Western blotting*

Western blots were performed as previously described[3]. Briefly, cells were lysed directly in pre-heated Laemmli buffer, quantified (BCA protein quantification kit – Pierce, Appleton WI, USA), loaded (20 µg) in standard 10% SDS-PAGE gels and transferred to nitrocellulose membranes (Bio-Rad, Hercules, CA, USA). Membranes were blocked with 5% skim milk for 30 min and incubated with primary antibodies (anti-p.STAT1 - 1:1000, Cell Signaling #9167; anti-FLAG - 1:1000, Sigma #F1804; anti-tubulin - 1:5000, Abcam #Ab18251) (Cell Signaling, Danves, MA, USA; Abcam, Cambridge, UK) diluted in 5% BSA in TBST buffer, overnight at 4˚C. Membranes were extensively washed, incubated in appropriate HRP-conjugated secondary antibodies (Sigma), washed, incubated with ECL Prime (Amersham, Amersham, UK) and the signal was detected using a Chemidoc MP Imaging System (Bio-Rad, Hercules, CA, USA). Signals were quantified using ImageJ software.

*Immunofluorescence staining for ADP-ribose and FLAG*

As previously described[3], A549 cells, either transduced with empty vector control or lentiviral constructs for FLAG-tagged SARS-CoV-2 macrodomain overexpression were seeded in microscopy-compatible plastic 96-well plates at the density of $10^4$ cells/well (Corning, Corning, NY, USA), treated as required, washed with PBS and fixed with 4% EM-grade PFA (EMS, Dormat, Switzerland), which was subsequently quenched twice with 0.1 M glycine and washed in PBS. After permeabilization in 0.2% TritonX-100 in PBS, samples were blocked in 10% FBS in permeabilization solution and incubated with primary antibodies (anti-ADP-ribose - 1:500, Millipore #MABE1016; anti-FLAG - 1:500, Sigma #F1804) (Millipore, Burlington MA, USA) for 1h at room temperature in blocking solution. Samples were extensively washed in PBS, incubated with appropriate fluorescently labelled secondary antibodies (Thermo), stained with DAPI (Thermo), washed and maintained in 30% glycerol. For Western blot the seeding density was $2 \times 10^5$ cells/well.

Fluorescence microscopy images were acquired on a customized TissueFAXS i-Fluo system (TissueGnostics, Wien, Austria) mounted on a Zeiss AxioObserver 7 microscope (Zeiss, Oberkochen, Germany), using 20x Plan-Neofluar (NA 0.5) objective and an ORCA Flash 4.0 v3 camera (Hamamatsu, Hamamatsu city, Japan). Acquired images were analyzed using StrataQuest software (TissueGnostics, Wien, Austria).

PAR immunofluorescence experiments were repeats were 8, for STAT1 phosphorylation or FLAG immunoblotting repeats were 4. All values were normalized to the IFNγ-treated empty vector control.

*SARS-CoV-2 spike protein binding assay*

Assessment of SARS-CoV2 RBD and hACE2 binding inhibition was performed using RayBio® COVID-19 spike-ACE2 binding assay kit (CoV-SACE2-1, RayBiotech Inc., https://www.raybiotech.com/covid-19-spike-ace2-binding-assay-kit-en/?variation_id=107121). Recombinant SARS-CoV-2 spike RBD protein coated wells were treated with the compounds (rucaparib, stenoparib, olaparib) in the range of concentration from 1 µM up to 500 µM, then incubated for 16 h at 4 °C. Next, the recombinant hACE2 protein was added and the protocol provided by the manufacturer was followed. In brief: unbound hACE2 protein was removed by washing, and binding was assessed based on HRP-conjugated IgG in the presence of 3,3',5,5'-tetramethylbenzidine (TMB) substrate. The HRP-conjugated IgG binds to the hACE2 protein and reacts with the TMB solution, producing a blue color that is proportional to the amount of bound hACE2. The HRP-TMB reaction is halted with the addition of the Stop Solution, resulting in a blue-to-yellow color change. The intensity of the yellow color is then measured by absorbance at 450 nm with a microplate reader SpectraMax® iD5 (Molecular Devices, LLC., San Jose, CA, USA). Data points were obtained as average of duplicates and $IC_{50}$-curves were fitted by non-linear regression using GraphPad Prism.

*Molecular modeling*

We have used the FTMap protein mapping algorithm to identify possible binding sites on the protein-protein interaction surface of the spike protein receptor-binding domain (RBD) vs. the human ACE2 receptor[5,6]. Briefly, the FTMap method distributes small organic probe molecules on a dense grid defined on the protein surface, finds the most favorable positions for each probe type, and identifies preferred binding hotspots as regions that bind multiple probe clusters. Here, we have used the experimentally determined structures of the ACE2-RBD complex for the B.1.5 ("Wuhan", PDB ID: 6M0J)[7], B.1.617.2 („Delta", PDB ID: 7WBQ) and B.1.1.529 („Omicron", PDB ID: 7WBP) variants[8], and observed exactly one binding hotspot at the protein-protein interaction surface of all three variants, characterized by the residues 493-498, with further sidechains R403, E406, Y449, Y453, N501 and Y505 in the vicinity. After preparing the structure of rucaparib with LigPrep[9], ligand docking was carried out with the single precision (SP) mode of Glide [10,11], and the binding mode was visualized with Maestro.

*Sequence homology alignment*

spike protein amino acid sequence was retrieved from https://viralzone.expasy.org/9556. Sequences were retrieved for the wild type (https://www.uniprot.org/uniprot/P0DTC2), the alpha (https://www.ncbi.nlm.nih.gov/protein/QWE88920), the beta (https://www.ncbi.nlm.nih.gov/protein/QRN78347), the gamma (https://www.ncbi.nlm.nih.gov/protein/QVE55289), the delta (https://www.ncbi.nlm.nih.gov/protein/QWK65230), the omicron BA.1 (https://www.ncbi.nlm.nih.gov/protein/UFO69279.1?report=fasta), the omicron BA.2 (https://www.ncbi.nlm.nih.gov/protein/UJE45220.1?report=fasta), and the epsilon (https://www.ncbi.nlm.nih.gov/protein/QQM19141 ) variants. The relevant parts of the sequence were aligned using the ClustalW algorithm (https://www.genome.jp/tools-bin/clustalw). All sites were accessed the 1st April 2022.

On the figure conserved amino acids as compared to the wild type variant are in green. The non-conserved amino acids are represented in red.

*Expression and purification of the receptor binding domain of the spike protein*

The receptor-binding domain (RBD) of the SARS-CoV-2 virus spike protein ($^{319}$Arg-$^{541}$Phe) (Wuhan-Hu-1) was used in the experiments. Suspension HEK-293 cells stably expressing the RBD secreted to the culture media were created by the Sleeping Beauty transposon method using the p10-IRES2-eGFP vector construct as in a different application, described previously [31].

The RDB sequence, as produced by Amanat et al. [32], was inserted to the Sleeping Beauty transposon plasmid. The sequence on the N-terminal includes a signal peptide responsible for the secretion of the protein to the culture media and 6x His tag was introduced on the Cterminal. Shaken cultures of suspension-adapted HEK293 cells were grown in a serum-free FreeStyle 293 expression medium (Gibco, Waltham, MA, USA, Cat. no. 12338018) incubated at 37°C and 5% CO2, shaken at 100 rpm. The cells were sub-cultured every 3-4 days with seeding densities of 2 ×105 cells/mL to promote cell growth and scale-up. The RBD was produced in a 300 mL shake flask cell culture. Cells were removed after 5-6 days by centrifugation (20 min, 1000 rpm, Rotanta 460R, Hettich, Kirchlengern, Germany). The purification of RBD was accomplished by immobilized nickel ion affinity chromatography. The supernatant was loaded into a 5 mL HisTrap HP column (GE Healthcare, Chicago, IL, USA), which was connected to an FPLC system (ÄKTA pureTM, GE Healthcare, Chicago, IL, USA). Protein bound to the column was eluted by imidazole (200 mM, pH 7.4). The protein-containing fraction was concentrated while the elution buffer was replaced by phosphate buffer (pH 6.8) using a centrifugal filter device. Centrifugation (Rotanta 460R, Hettich, Kirchlengern, Germany) was performed at 4000 g for 20 minutes using a 30 KDa MWCO filter (Vivacell 100, Sartorious, Göttingen, Germany). The purity of the produced protein was confirmed by SDS-PAGE and the protein concentration was determined by UV-Vis spectroscopy at 280 nm.

*Nuclear Magnetic Resonance (NMR)-based investigation of spike-rucaparib interaction*

Solution phase NMR experiments were performed under the following conditions.

The RBD of spike (~ 28 kDa) + rucaparib camsylate assay mixture contained 33 μM RBD (in 540 μL) and 1 mM rucaparib (dissolved in DMSO-$d_6$ in 20 μL) + 10% $D_2O$ for lock in sodium-phosphate buffer (0.2 M, pH 6.8). Rucaparib was in ~30 fold excess. The RBD and rucaparib were assessed also without the other partner under the same conditions as references.

Bruker Avance Neo 700 MHz spectrometer (Bruker, Billerica, MA, USA) equipped with a Prodigy TCI cryoprobe was used. Experiments were run at 298K temperature and 10% $D_2O$ was added to the solution for stabilizing the magnetic field via the $^2$H lock channel. Typical 90 degree $^1$H pulse duration was 11.5 μs. The residual DMSO-$d_6$ signal (2.59 ppm) was used as $^1$H chemical shift reference. The NMR verification of the structure of rucaparib-camphor sulfonic acid (camsylate) salt was carried out in pure DMSO-$d_6$ solvent and yielded a full assignment of all $^1$H and $^{13}$C signals (spectra not shown). The assignments of aromatic (H15, H16, H18,H19) and olefinic (H11, H13) protons could be easily transferred to the buffer, containing rucaparib with or without the RBD protein. Excitation sculpting water suppression $^1$H-NMR spectra using manufacturers 'zgesfpgp' pulse sequence was applied (**FigS1G**)[12]. The drug ligand signal appear as sharp signals while the protein signals are weak and broad (due to concentration and MW differences). A relatively strong and well separated RBD signal at 0.4 ppm was selected for selective irradiation in STD-NMR (saturation transfer difference) experiments (weaker RBD signals below 0 ppm were also tested, and gave observable, but weaker responses)[13-16]. For STD we used manufacturers 'stddiffgp19.3' pulse sequence with watergate water suppression[17]. Selective saturation was achieved using a series of 50 ms selective 90 degree Gaussian pulses, resulting in 2000 ms total irradiation time. The reference experiment was carried out identically, except that an off resonance irradiation at -40 ppm was applied. A total of 12160 scans interrupted with 2s relaxation delays were coadded in both the on-resonance and the off-resonance (reference) experiments yielding one day total experiment time. In the difference spectrum of the two experiments (**FigS1H**) the irradiated broad RBD signal is the strongest among protein signals, while the six protons of rucaparib ligand-probably closest to the binding site- gave the strongest responses. In the control experiment with no RBD protein, the Rucaparib did not show the characteristic difference signals. An independent corroboration of Rucaparib binding to RBD was carried out with the (non-selective) transferred NOESY[18] experiment ('noesygpph19' pulse sequence). Again, the fingerprint signals gave cross-peaks with the same sign relative to the diagonal in the 2D NOESY map while they were missing in a sample without the protein.

*Transcription factor activation assay*

Human peripheral blood mononuclear cells were differentiated to macrophages as described above. Cells were induced with the spike protein of SARS-CoV-2 (20 nM, 24 hours) and were treated with rucaparib in the concentrations indicated. Cell were then scraped whole cell lysates were used to perform transcription factor binding assays using TransAm kits (ActiveMotif Tegernheim, Bayern, Germany) similar to[19]. Stat family (Cat. No. 42296) was used in the study.

*Statistical analyses*

All experiments were repeated on at least three separate occasions, often with multiple parallel replicates processed on the same day, but treated as independently as possible. All graphs and statistical analyses were generated using GraphPad Prism v.8.0.1 software.

Numerical values are presented as the average ± SD. Normality was checked. . *, **, *** represent statistically significant differences between the controls/vehicle-treated cells and patients/treated cells at $p<0.05$, $p<0.01$ and $p<0.001$, respectively.

Two-tailed Student's t-test on Figure 1 panel C and D

One-way ANOVA was used on Figure 1 panel B, Supplementary Figure 1 panels A, C, E and F, in the post-hoc test all treatments were compared to the control.

One-way ANOVA was used on Figure 2 E and F and throughout Supplementary Figure 2, followed by a post-hoc test comparing all samples to the LPS or spike-treated groups.

**Captions to supplementary Figures**

**Supplementary Figure 1. Olaparib and talazoparib do not block SARS-CoV-2 infection and rucaparib does not inhibit the macrodomain of SARS-CoV-2**

**(A)** The antiproliferative and toxic effects of olaparib and talazoparib were tested in Vero E6 cells infected with of B.1.5 variant SARS-CoV-2. **(B)** Predicted binding mode of rucaparib (green) at the protein-protein binding surface of the SARS-CoV2 RBD (fawn), overlaid on the position of the ACE2 receptor (transparent blue). **(C-F)** A549 cells, either transduced with empty vector control (e.v.) or lentiviral constructs for FLAG-tagged SARS-CoV-2 macrodomain overexpression (MD) were treated with either vehicle or different combinations of 100U IFNγ, 500 nM or 20 μM rucaparib for 24 hours, as shown. ADP-ribosylation was detected by immunofluorescence (C) and STAT1 phosphorylation, FLAG or tubulin levels were determined by immunoblotting (D-F). All values were normalized to the IFNγ-treated e.v. control. **(G)** Excitation sculpting water suppression $^{1}$H-NMR spectra using the manufacturer's 'zgesfpgp' pulse sequence. Blue: no RBD, red: with RBD. **(H)** Whole difference spectrum of the Rucaparib alone (blue), Rucaparib with RBD (green) and buffer alone (red). The red rectangle marks the region displayed on **Fig.2B**.

**Supplementary Figure 2. Rucaparib inhibits the LPS-induced overexpression of IL-8 and IL-10**

**(A)** Human primary monocyte-derived macrophages were challenged with LPS (100 ng/mL, 24 hours) and indicated interleukins were determined. Three individual donors are displayed. **(B)** A549 cells were induced with poly(I:C) and STAT1 phosphorylation was determined as described in Materials and Methods. **(C)** Stat1, 3, 5A and 5B activation was determined in human macrophages using the TransAM kit as described in the Materials and methods. One donor is displayed.

**Supplementary Table 1. Patient characteristics**

For comparing age, normality was checked and two-sided, non-paired t-test was applied. AVG – average, CVE – cardiovascular event, DCM – dilated cardiomyopathy, F – female, IHD – ischaemic heart disease, GERD - Gastroesophageal reflux disease, M – male, n.s. – not significant

| | **Control patients** | | | **COVID-19 patients** | | |
|---|---|---|---|---|---|---|
| | **Patient identifier** | **Sex** | **Age** | **Patient identifier** | **Sex** | **Age** |
| | 1 | F | 66-70 | 1 | M | 56-60 |
| | 2 | F | 86-90 | 2 | F | 86-90 |
| | 3 | F | 81-85 | 3 | F | 81-85 |
| | 4 | M | 86-90 | 4 | F | 61-65 |
| | 5 | M | 66-70 | 5 | F | 81-85 |
| | 6 | F | 86-90 | 6 | M | 56-60 |
| | 7 | M | 56-60 | 7 | M | 61-65 |
| | 8 | F | 76-80 | 8 | F | 56-60 |
| | 9 | M | 81-85 | 9 | F | 76-80 |
| | 10 | M | 65-70 | 10 | M | 61-65 |
| | | | | 11 | M | 61-65 |
| | | | | 12 | M | 76-80 |
| | | | | 13 | F | 71-75 |
| | | | | 14 | M | 61-65 |
| | | | | 15 | M | 61-65 |
| **AVG** | 10 patients | 50% M 50% F | 77.1 ± 10.5 yrs | 15 patients | 53,3% M 46,7% F | 69 ± 10.9 yrs$^{ns}$ |

**Supplementary Table 2. Conditions for immunohistochemistry**

Abbreviation: CC1 - ULTRA Cell Conditioning Solution (Ventana Medical Systems, Oro Valley, AZ, USA)

| Primary Antibody | | | | | Antigen retrieval: HIER | | Immunohistochemical stainers | | | |
|---|---|---|---|---|---|---|---|---|---|---|
| **Antibody name** | **Vendor** | **Host** | **Dilution factor** | **Incubation time/temp.** | **Method** | **Time/Temp.** | **Visualization system** | **Enhancement** | **Counterstain** | **Staining Platform** |
| **Anti-4 Hydroxynonenal antibody (ab46545)** | **Abcam** | rabbit polyclonal | 1/400 | 32'/37 °C | CC1, pH8,5 | 36' ',95°C; | UltraView Universal DAB Detection Kit | CuSO4 | Hematoxylin II | VENTANA BenchMark ULTRA |
| **Poly(ADP-ribose) monoclonal antibody (10H). (ALX-804-220-R100)** | **Enzo** | mouse monoclonal | 1/2000 | 32'/37 °C | | 64 ',95°C; | | | | |
| **SARS-CoV-2 spike Glycoprotein S1 ab/ (ab275759)** | **Abcam** | rabbit polyclonal | 1/1000 | 1h /37 °C | | 56 ',100°C; | | | | |
